# Post-vaccination SARS-CoV-2 infections and incidence of the B.1.427/B.1.429 variant among healthcare personnel at a northern California academic medical center

**DOI:** 10.1101/2021.04.14.21255431

**Authors:** Karen B. Jacobson, Benjamin A. Pinsky, Maria E. Montez Rath, Hannah Wang, Jacob A. Miller, Mehdi Skhiri, John Shepard, Roshni Mathew, Grace Lee, Bryan Bohman, Julie Parsonnet, Marisa Holubar

**Affiliations:** Department of Medicine, Division of Infectious Diseases and Geographic Medicine, Stanford University School of Medicine, Stanford, CA, USA; Department of Pathology, Stanford University School of Medicine, Stanford, CA, USA; Department of Medicine, Division of Nephrology, Stanford University School of Medicine, Stanford, CA, USA; Department of Radiation Oncology, Stanford University School of Medicine, Stanford, CA, USA; Department of Medicine, Primary Care and Population Health, Stanford University School of Medicine, Stanford, CA, USA; Department of Quality, Patient Safety and Clinical Effectiveness, Stanford Health Care, Stanford, CA, USA; Department of Pediatrics, Division of Infectious Diseases, Stanford University School of Medicine, Stanford, CA, USA; Workforce Health and Wellness, Stanford University School of Medicine, Stanford, CA, USA; Department of Epidemiology and Population Health, Stanford University School of Medicine, Stanford, CA, USA

## Abstract

**Background:** Distribution of mRNA-based SARS-CoV-2 vaccines to healthcare personnel (HCP) in the United States began in December 2020, with efficacy ≥ 90%. However, breakthrough infections in fully vaccinated individuals have been reported. Meanwhile, multiple SARS-CoV-2 variants of concern have emerged worldwide, including the B.1.427/B.1.429 variant first described in California. Little is known about the real-world effectiveness of the mRNA-based SARS-CoV-2 vaccines against novel variants including B.1.427/B.1.429.

**Methods:** In this quality improvement project, post-vaccine SARS-CoV-2 cases (PVSCs) were defined as individuals with positive SARS-CoV-2 nucleic acid amplification test (NAAT) after receiving at least one dose of a SARS-CoV-2 vaccine. Chart extraction of demographic and clinical information was performed, and available specimens meeting cycle threshold value criteria were tested for L452R, N501Y and E484K mutations by RT-PCR.

**Results:** From December 2020 to March 2021, 189 PVSCs were identified out of 22,729 healthcare personnel who received at least one dose of an mRNA-based SARS-CoV-2 vaccine. Of these, 114 (60.3%) occurred within 14 days of first vaccine dose (early post-vaccination), 49 (25.9%) within 14 days of the second vaccine dose (partially vaccinated), and 26 (13.8%) ≥14 days after the second dose (fully vaccinated). Of 115 samples available for mutation testing, 42 were positive for L452R alone, presumptive of B.1.427/B.1.429; three had N501Y mutation alone and none were found with E484K mutation. Though on univariate analysis partially- and fully-vaccinated PVSCs were more likely than early post-vaccination PVSCs to be infected with presumptive B.1.427/B.1.429, when adjusted for community prevalence of B.1.427/B.1.429 at the time of infection, partially- and fully-vaccinated PVSC did not have statistically significantly elevated risk ratios for infection with this variant (RR 1.40, 95% CI 0.81-2.43 and RR 1.13, 95% CI 0.59-2.16, respectively).

**Conclusions:** The great majority of PVSCs occurred prior to the expected onset of full, vaccine-derived immunity. Although the B.1.427/B.1.429 variant did not represent a significantly higher proportion of PVSCs than expected, numbers were small and there was a trend towards higher representation in the partially- and fully-vaccinated subset. Continued infection control measures in the workplace and in the community including social distancing and masking, particularly in the early days post-vaccination, as well as continued variant surveillance in PVSCs, is imperative in order to anticipate and control future surges of infection.

## Introduction

SARS-CoV-2 has infected 120 million people worldwide, causing at least 2.6 million deaths from COVID-19 since December 2019.(1) To control the pandemic, vaccines have been developed at unprecedented rates, with mRNA-based vaccines developed by Pfizer and Moderna obtaining emergency use authorization (EUA) approval in the United States in December 2020. Vaccine administration to elderly individuals, healthcare workers and other first responders began shortly thereafter. In clinical trials and subsequent observational studies after vaccine rollout, both mRNA vaccines proved effective in preventing symptomatic and severe disease.(2-4) In clinical trials, the vaccines demonstrated 52-95% efficacy against symptomatic disease 14 days after the first dose, and 95% efficacy seven days after the second dose.(3, 4) Early reports of real-world experience among vaccinated health care personnel suggest 80% effectiveness >14 days after first dose and 90% >14 days after second dose.(5-7) Emerging evidence indicates both vaccines may also be effective in preventing asymptomatic infections(8) that can lead to ongoing transmission.

The hope for a rapid end to the COVID-19 pandemic through effective vaccination has been dampened by the emergence of numerous SARS-CoV-2 variants that have appeared worldwide, including variants originating in the UK (B.1.1.7), South Africa (B.1.351), Brazil (P.1), and California (B.1.427/B.1.429).(9) The B.1.427/B.1.429 variant, which carries a L452R substitution mutation in the spike protein(10), was first detected in October 2020 and by January 2021 accounted for 35% of all SARS-CoV-2 cases detected in California.(9) Evidence of reduced neutralizing activity of naturally-, vaccine-acquired and monoclonal antibodies against these variants including B.1.427/B.1.429(11-16), and possibly increased transmissibility(10, 17) and virulence, is of great concern. Humoral immunity, however, is not the only factor in determining protection against infection(18) and more real-world data is needed to determine effectiveness of the available SARS-CoV-2 vaccines against SARS-CoV-2 and its variants.

Stanford Health Care (SHC) began vaccinating healthcare personnel (HCP) against SARS-CoV-2 on December 18, 2020, during a surge in COVID-19 cases when the B.1.427/B.1.429 variant was rapidly spreading.(9) We performed a retrospective quality improvement project of post-vaccine SARS-CoV-2 cases (PVSCs) to help inform future guidance on infection control recommendations for vaccinated HCP. Our aims were to: 1) define and characterize post-vaccine SARS-CoV-2 infections in HCP, 2) evaluate our HCP vaccination program, and 3) determine the role of variants in causing PVSCs.

## Methods

On December 18, 2020, SHC began vaccinating HCP with Pfizer mRNA SARS-CoV-2 vaccine, initially to patient-facing frontline HCP in high acuity settings, then expanding to all patient-facing frontline HCP on December 28, and finally to all non-patient facing medical staff members and health system employees beginning January 8, 2021. Moderna mRNA vaccine was also available as of January 22, 2021. All HCP were required to self-monitor for COVID-related symptoms and, if they developed, have same-day assessment and testing. Some HCP also participated in voluntary weekly asymptomatic NAAT testing.

### Data Collection

Post-vaccine SARS-CoV-2 cases (PVSCs) were defined as individuals identified by occupational health case notification and chart extraction with a positive SARS-CoV-2 nucleic acid amplification testing (NAAT) after receiving at least one dose of a SARS-CoV-2 vaccine. Initial respiratory SARS-CoV-2 NAAT was conducted on a variety of platforms(19-21) including: 1) a previously-described laboratory-developed reverse transcription quantitative polymerase chain reaction (RT-qPCR) targeting the envelope gene (*E* gene) on the Rotor-Gene Q (Qiagen, Germantown, MD); 2) a laboratory-developed RT-qPCR assay utilizing a PerkinElmer kit targeting the ORF1ab and nucleocapsid gene ([*N* gene] PerkinElmer, San Jose, CA); 3) Panther Fusion SARS-CoV-2 (Hologic, Marlborough, MA), a high-throughput RT-qPCR method targeting open reading frame 1ab (ORF1ab); 4) Aptima SARS-CoV-2 (Panther System, Hologic), a transcription mediated amplification method targeting ORF1ab; 5) GeneXpert Xpress SARS-CoV-2 (Cepheid, Sunnyvale, CA), a rapid RT-qPCR method targeting both *E* and *N* genes; 6) cobas Liat SARS-CoV-2 & Influenza A/B (Roche, Indianapolis, IN), a point-of-care RT-PCR method targeting ORF1ab and *N* gene; 7) e-Plex SARS-CoV-2 (Genmark, Carlsbad, CA), a rapid RT-PCR method targeting the *N* gene.

Chart extraction data included age and gender, dates of reported vaccine doses, date of positive SARS-CoV-2 NAAT, presence of symptoms of COVID-19 and date of symptom onset, household exposures to SARS-CoV-2, previous history of a COVID-19 diagnosis prior to vaccination, and immunocompromising conditions or medications. Occupational health records were also used to quantify the total number of vaccinations administered to HCP, total number of SARS-CoV-2 tests performed, and total number of positive SARS-CoV-2 tests that occurred during the study period.

Between December 1, 2020 and March 1, 2021 all available specimens testing positive for SARS-CoV-2 by NAAT with RT-qPCR cycle threshold (C_t_) ≤ 30 or transcription-mediated amplification relative light units (RLU) ≥ 1,100 during this period were subject to multiplex allele-specific genotyping RT-qPCR targeting three spike mutations associated with known variants of concern, including N501Y (B.1.1.7, B.1.351, P.1), E484K (B.1.351, P.1), and L452R (B.1.427/B.1.429).(22) The screening threshold was changed to C_t_ ≤ 34 for specimens collected March 1^st^ and onwards. This variant data was then used to compared incidence of variant-associated mutations in PVSCs with that of the general population.

Data collection for this quality improvement project was approved by hospital privacy compliance and deemed to not be human subjects research by the Stanford University School of Medicine Panel on Human Subjects in Medical Research.

### Exposures and outcomes

The main exposure was vaccination status at time of positive test, defined as early post-vaccination (positive test ≤14 days after first vaccine dose), partially vaccinated (positive test >14 days after first vaccine dose to ≤14 days after the second vaccine dose), and fully vaccinated (positive test >14 days after second vaccine dose).

The primary outcome was the presence of L452R mutation consistent with presumptive B.1.427/B.1.429.(22) If no L452R, N501Y, or E484K mutations were identified, the infection was designated as wild-type SARS-CoV-2.

### Statistical analysis

Statistical analyses were performed in SPSS Version 27. Chi square, Fisher’s exact test, independent samples T tests, and a modified Poisson regression model (log-Poisson generalized linear model with robust standard errors)(23) was used to compare individuals with and without presumptive B.1.427/B.1.429 and who tested positive ≤ 14 or >14 days since their first and second vaccine doses.

## Results

Between December 18, 2020 and March 15, 2021, 22,729 HCP received at least one dose of Pfizer (n=20,559) or Moderna (n=2,170); by March 15, 2021, 22,053 HCP had completed the 2-shot series. In that same period, 40,041 SARS-CoV-2 tests were performed among HCP within the SHC system of which 394 (1.0%) were positive for SARS-CoV-2 by NAAT. From December 19, 2020 to April 2, 2021, 189 PVSCs were identified, representing 0.8% of all SHC employees who received at least one dose of an mRNA vaccine. Of these 189 PVSCs, 173 (91.5%) received at least one dose of the Pfizer mRNA vaccine and 15 (7.9%) received at least one dose of the Moderna mRNA vaccine; one PVSC vaccinated outside of the SHC system had no documentation of vaccine brand.

The median age of PVSCs was 38 years (IQR 32-48) and 125 (66.1%) were female; 42 (22.2%) were nurses (Table 1). Few individuals had immunocompromising conditions (n=7, 3.7%). Seventy-six (41.8%) reported a known household contact with SARS-CoV-2 infection at the time of their positive SARS-CoV-2 NAAT.

**Table 1.**
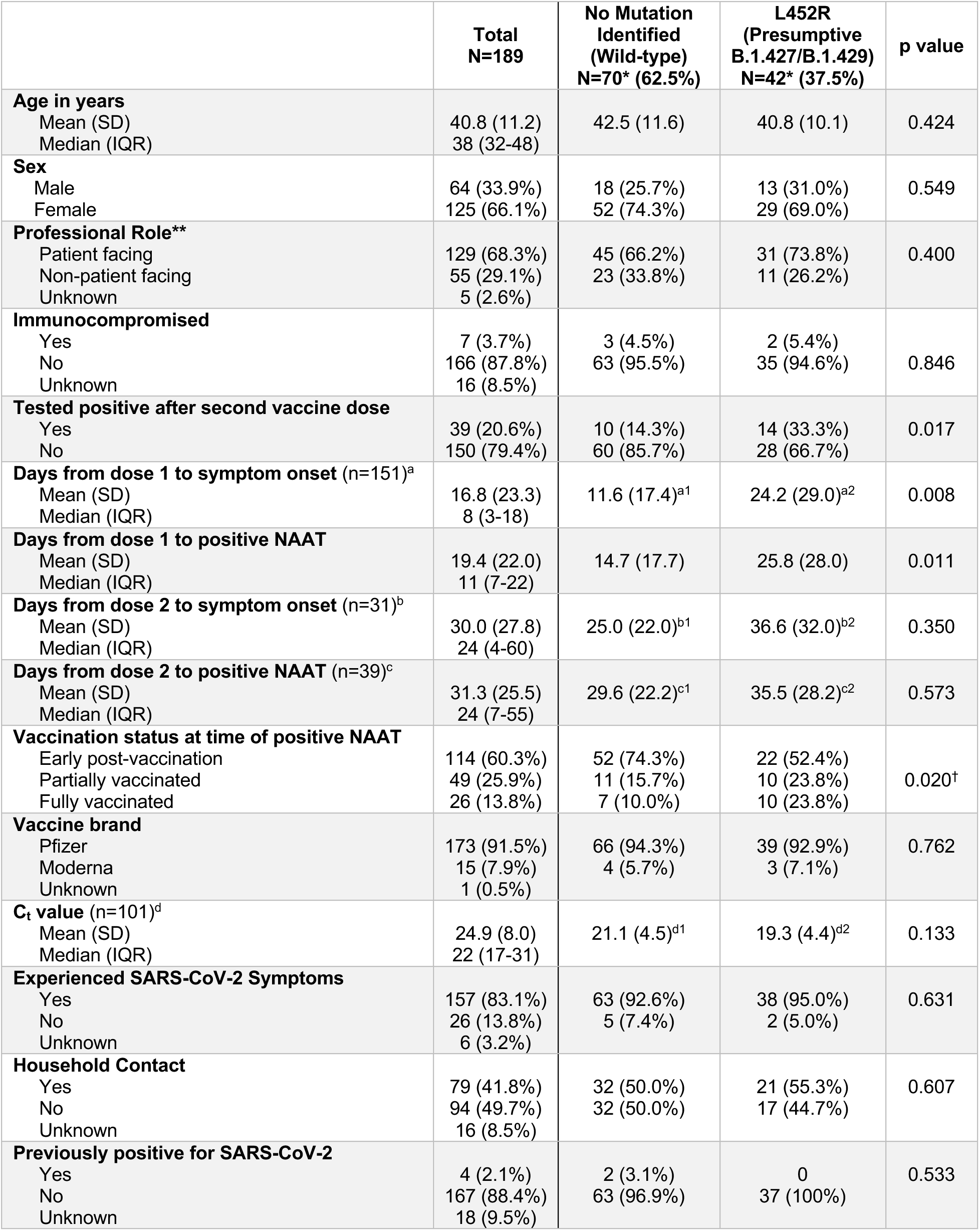
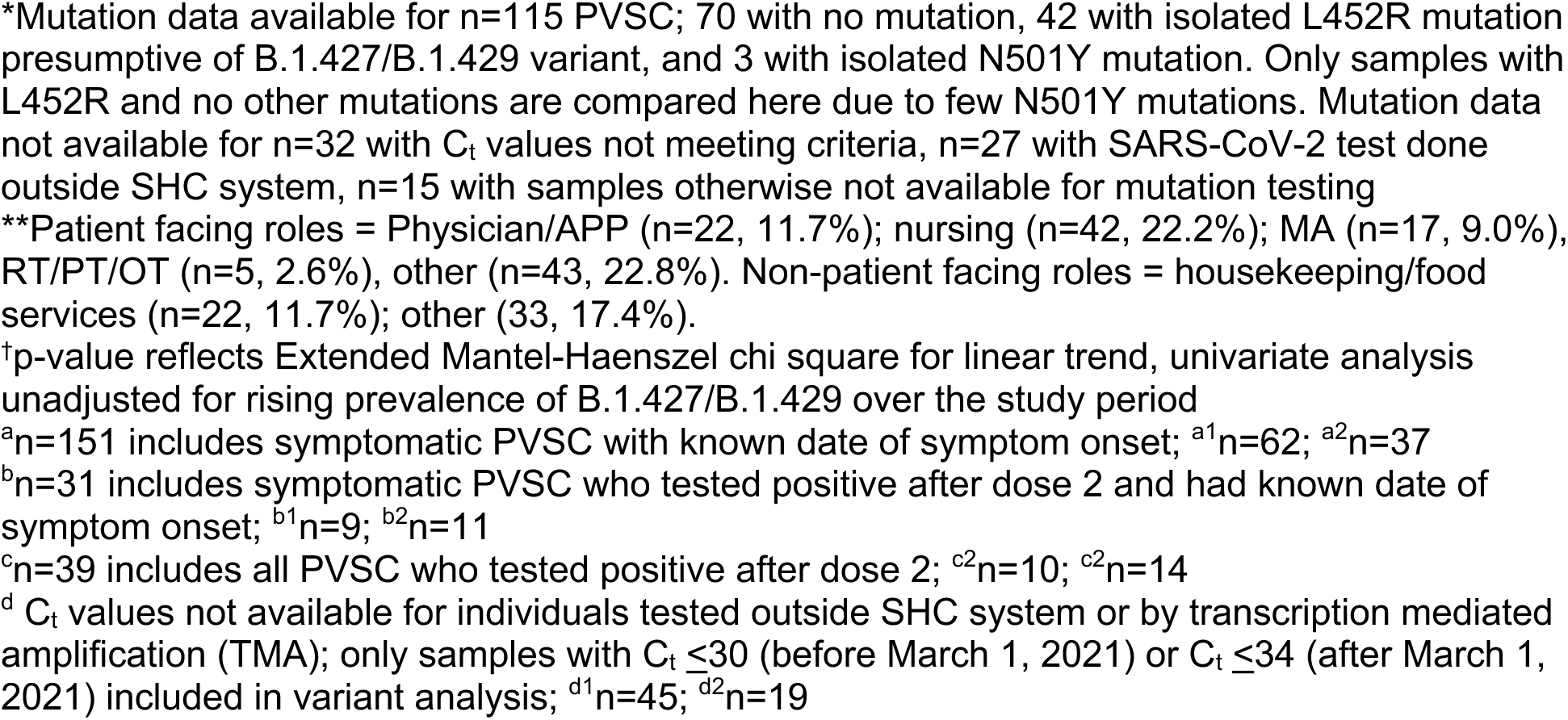
Characteristics of post-vaccine SARS-CoV-2 cases among healthcare personnel.

|  | **Total**<br>**N=189** | **No Mutation**<br>**Identified**<br>**(Wild-type)**<br>**N=70\* (62.5%)** | **L452R**<br>**(Presumptive**<br>**B.1.427/B.1.429)**<br>**N=42\* (37.5%)** | **p value** |
| --- | --- | --- | --- | --- |
| **Age in years** |  |  |  |  |
| Mean (SD) | 40.8 (11.2) | 42.5 (11.6) | 40.8 (10.1) | 0.424 |
| Median (IQR) | 38 (32-48) |  |  |  |
| **Sex** |  |  |  |  |
| Male | 64 (33.9%) | 18 (25.7%) | 13 (31.0%) | 0.549 |
| Female | 125 (66.1%) | 52 (74.3%) | 29 (69.0%) |  |
| **Professional Role\*\*** |  |  |  |  |
| Patient facing | 129 (68.3%) | 45 (66.2%) | 31 (73.8%) | 0.400 |
| Non-patient facing | 55 (29.1%) | 23 (33.8%) | 11 (26.2%) |  |
| Unknown | 5 (2.6%) |  |  |  |
| **Immunocompromised** |  |  |  |  |
| Yes | 7 (3.7%) | 3 (4.5%) | 2 (5.4%) | 0.846 |
| No | 166 (87.8%) | 63 (95.5%) | 35 (94.6%) |  |
| Unknown | 16 (8.5%) |  |  |  |
| **Tested positive after second vaccine dose** |  |  |  |  |
| Yes | 39 (20.6%) | 10 (14.3%) | 14 (33.3%) | 0.017 |
| No | 150 (79.4%) | 60 (85.7%) | 28 (66.7%) |  |
| **Days from dose 1 to symptom onset (n=151)<sup>a</sup>** |  |  |  |  |
| Mean (SD) | 16.8 (23.3) | 11.6 (17.4)<sup>a1</sup> | 24.2 (29.0)<sup>a2</sup> | 0.008 |
| Median (IQR) | 8 (3-18) |  |  |  |
| **Days from dose 1 to positive NAAT** |  |  |  |  |
| Mean (SD) | 19.4 (22.0) | 14.7 (17.7) | 25.8 (28.0) | 0.011 |
| Median (IQR) | 11 (7-22) |  |  |  |
| **Days from dose 2 to symptom onset (n=31)<sup>b</sup>** |  |  |  |  |
| Mean (SD) | 30.0 (27.8) | 25.0 (22.0)<sup>b1</sup> | 36.6 (32.0)<sup>b2</sup> | 0.350 |
| Median (IQR) | 24 (4-60) |  |  |  |
| **Days from dose 2 to positive NAAT (n=39)<sup>c</sup>** |  |  |  |  |
| Mean (SD) | 31.3 (25.5) | 29.6 (22.2)<sup>c1</sup> | 35.5 (28.2)<sup>c2</sup> | 0.573 |
| Median (IQR) | 24 (7-55) |  |  |  |
| **Vaccination status at time of positive NAAT** |  |  |  |  |
| Early post-vaccination | 114 (60.3%) | 52 (74.3%) | 22 (52.4%) | 0.020<sup>†</sup> |
| Partially vaccinated | 49 (25.9%) | 11 (15.7%) | 10 (23.8%) |  |
| Fully vaccinated | 26 (13.8%) | 7 (10.0%) | 10 (23.8%) |  |
| **Vaccine brand** |  |  |  |  |
| Pfizer | 173 (91.5%) | 66 (94.3%) | 39 (92.9%) | 0.762 |
| Moderna | 15 (7.9%) | 4 (5.7%) | 3 (7.1%) |  |
| Unknown | 1 (0.5%) |  |  |  |
| **C<sub>t</sub> value (n=101)<sup>d</sup>** |  |  |  |  |
| Mean (SD) | 24.9 (8.0) | 21.1 (4.5)<sup>d1</sup> | 19.3 (4.4)<sup>d2</sup> | 0.133 |
| Median (IQR) | 22 (17-31) |  |  |  |
| **Experienced SARS-CoV-2 Symptoms** |  |  |  |  |
| Yes | 157 (83.1%) | 63 (92.6%) | 38 (95.0%) | 0.631 |
| No | 26 (13.8%) | 5 (7.4%) | 2 (5.0%) |  |
| Unknown | 6 (3.2%) |  |  |  |
| **Household Contact** |  |  |  |  |
| Yes | 79 (41.8%) | 32 (50.0%) | 21 (55.3%) | 0.607 |
| No | 94 (49.7%) | 32 (50.0%) | 17 (44.7%) |  |
| Unknown | 16 (8.5%) |  |  |  |
| **Previously positive for SARS-CoV-2** |  |  |  |  |
| Yes | 4 (2.1%) | 2 (3.1%) | 0 | 0.533 |
| No | 167 (88.4%) | 63 (96.9%) | 37 (100%) |  |
| Unknown | 18 (9.5%) |  |  |  |

\*Mutation data available for n=115 PVSC; 70 with no mutation, 42 with isolated L452R mutation presumptive of B.1.427/B.1.429 variant, and 3 with isolated N501Y mutation. Only samples with L452R and no other mutations are compared here due to few N501Y mutations. Mutation data not available for n=32 with C<sub>t</sub> values not meeting criteria, n=27 with SARS-CoV-2 test done outside SHC system, n=15 with samples otherwise not available for mutation testing
\*\*Patient facing roles = Physician/APP (n=22, 11.7%); nursing (n=42, 22.2%); MA (n=17, 9.0%), RT/PT/OT (n=5, 2.6%), other (n=43, 22.8%). Non-patient facing roles = housekeeping/food services (n=22, 11.7%); other (33, 17.4%).
<sup>†</sup>p-value reflects Extended Mantel-Haenszel chi square for linear trend, univariate analysis unadjusted for rising prevalence of B.1.427/B.1.429 over the study period
<sup>a</sup>n=151 includes symptomatic PVSC with known date of symptom onset; <sup>a1</sup>n=62; <sup>a2</sup>n=37
<sup>b</sup>n=31 includes symptomatic PVSC who tested positive after dose 2 and had known date of symptom onset; <sup>b1</sup>n=9; <sup>b2</sup>n=11
<sup>c</sup>n=39 includes all PVSC who tested positive after dose 2; <sup>c2</sup>n=10; <sup>c2</sup>n=14
<sup>d</sup> C<sub>t</sub> values not available for individuals tested outside SHC system or by transcription mediated amplification (TMA); only samples with C<sub>t</sub> ≤30 (before March 1, 2021) or C<sub>t</sub> ≤34 (after March 1, 2021) included in variant analysis; <sup>d1</sup>n=45; <sup>d2</sup>n=19

The majority (n=114, 60.3%) of PVSCs occurred early post-vaccination (positive test ≤14 days after first vaccine dose) (Figure 1A); 49 (25.9%) occurred while partially vaccinated (positive test >14 days after first vaccine dose to ≤14 days after the second vaccine dose), and 26 (13.8%) occurred > 14 days after the second dose (Figure 1C). Of 157 PVSCs who experienced symptoms, 151 had known time of symptom onset; of these, 104 (68.9%) developed symptoms within 14 days of the first vaccine dose (Figure 1B). Fully vaccinated PVSCs who tested positive >14 days after the second vaccine dose had higher mean C_t_ values (30.5 vs 24.3, p=0.035, of n=101 individuals with known C_t_ value, Table 2).

**Table 2.** Age and C_t_ value by vaccination status.

|  | **Early post-vaccination**<br>**N=114 (60.3%)** | **Partially+Fully vaccinated**<br>**N=75 (39.7%)** | **P value** | **Early post-vaccination +Partially vaccinated**<br>**N=163 (86.2%)** | **Fully vaccinated**<br>**N=26 (13.8%)** | **P value** |
| --- | --- | --- | --- | --- | --- | --- |
| **Age in years**<br>Mean (SD) | 39.8 (10.8) | 42.3 (11.8) | 0.139 | 41.0 (11.5) | 39.1 (9.5) | 0.549 |
| **C<sub>t</sub> value (n=98)**<br>Mean (SD) | 22.8 (7.1) | 29.0 (8.2) | <0.001 | 24.3 (8.0) | 30.5 (7.2) | 0.035 |

T tests comparing age and C<sub>t</sub> value by vaccination status. Early post-vaccination = Positive SARS-CoV-2 NAAT ≤ 14 days from vaccine dose 1; Partially vaccinated = >14 days from vaccine dose 1 and ≤ 14 days from vaccine dose 2; Fully vaccinated = >14 days from vaccine dose 2.

**Figure 1.**
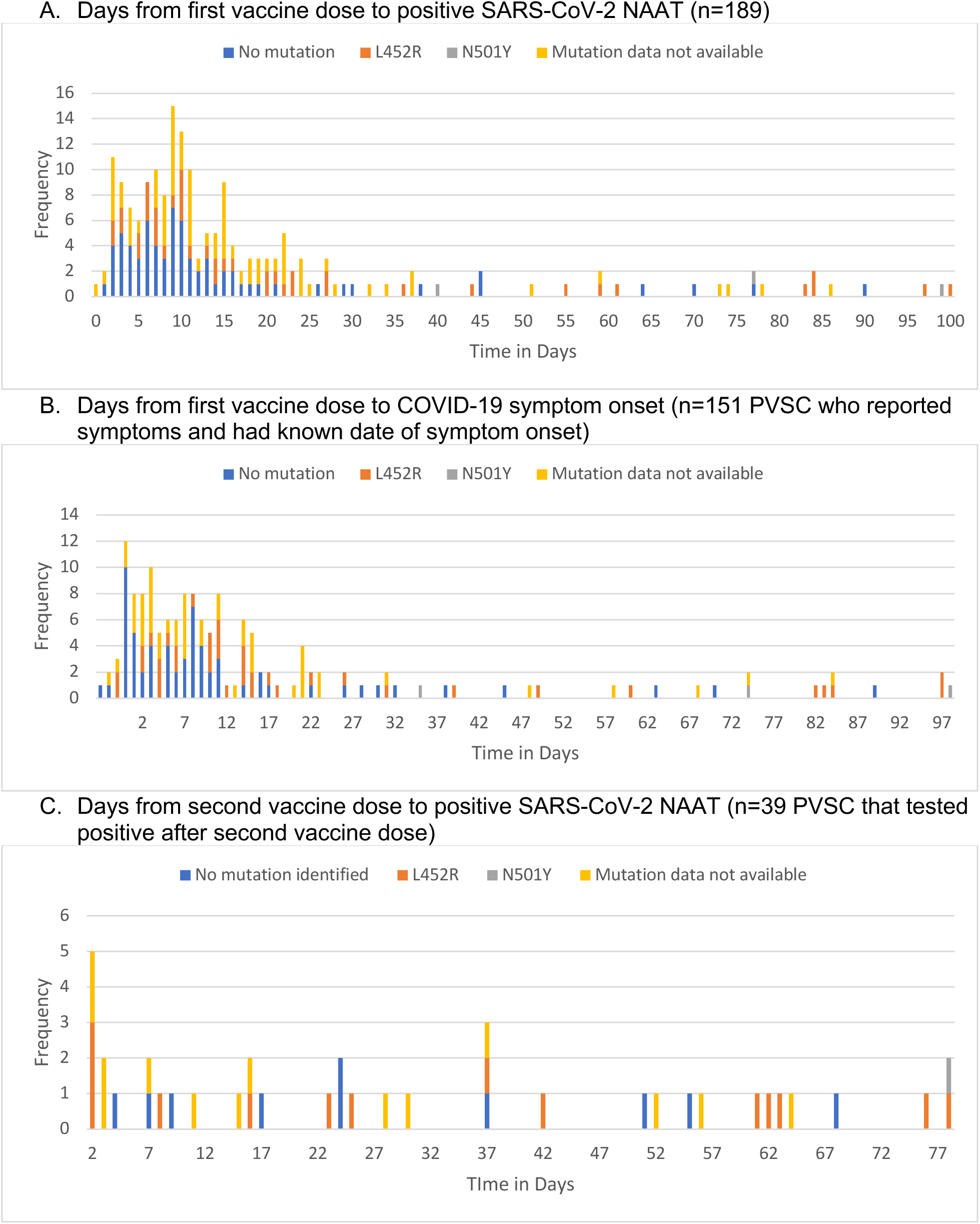
Time from vaccination to COVID-19 symptom onset and positive SARS-CoV-2 NAAT. Frequency of time from vaccination (Figure 1A=dose 1, Figure 1C=dose 2) or symptom onset (Figure 1B) to positive SARS-CoV-2 nucleic acid amplification test (NAAT) by presence of variant-associated mutation among post-vaccination SARS-CoV-2 cases. Presence of L452R mutation indicates presumptive B.1.427/B.1.429 variant.

The majority of PVSCs (n=140, 74.1%) identified to date were initially vaccinated in December 2020. The greatest number of post-vaccination positive tests occurred in individuals who had been vaccinated on December 22-23, 2020 (n=46, 24.3%), corresponding with the highest daily administration of vaccines (1940 doses given on 12/22/20 and 1934 given on 12/23/20). Of the PVSCs vaccinated on these days, 21 (45.7%) tested positive >14 days after their vaccination. The majority of positive tests in PVSCs also occurred in December 2020 and early January 2021, when most PVSCs were ≤14 days from their first vaccine dose, and also corresponding with the winter surge in SARS-CoV-2 cases in northern California. Over the study period, the B.1.427/B.1.429 variant rose in prevalence from representing 16.3% of SARS-CoV-2 cases detected by Stanford laboratory in mid-December 2020 to 51.0% of cases by late March 2021.(24)

The great majority (n=157, 83.1%) of PVSCs experienced symptoms of COVID-19 that began a median of 8 days (IQR 3-18 days) after the first vaccination dose. Eighteen PVSCs had developed symptoms of COVID-19 prior to or on the day of their first vaccine dose. Two PVSCs—both of whom tested positive <14 days after their first vaccine dose—were hospitalized with wild-type virus but there were no deaths. Of the 26 patients with no symptoms, 7 PVSCs (26.9%) had at least one repeat negative SARS-CoV-2 NAAT within 48 hours of their positive test. PVSCs who reported symptoms consistent with COVID-19 had lower mean C_t_ value (24.2 vs 30.9, p=0.011) than those without symptoms but did not differ in terms of age or time from first vaccine dose to positive test (data not shown).

Four PVSCs (2.2%) had previous documentation of SARS-CoV-2 infection between 22-98 days before their vaccinations; two reported symptoms at the time of their positive post-vaccination test. Two of these individuals had specimens available for mutation screening, and neither had evidence of infection with a variant of concern.

Tests for novel variant-associated mutations were performed on samples from 115 (60.8%) PVSCs; samples were unavailable for testing from 27 (14.3%) who were tested outside the SHC system, 32 (16.9%) did not meet C_t_ value criteria, and 15 (7.9%) that were discarded or otherwise unavailable. Three PVSCs were found to have N501Y mutation; all were symptomatic, two tested positive while partially vaccinated (>14 days after first vaccine dose but prior to the second dose) and one was fully vaccinated (>14 days after second dose). No E484K mutations were found by RT-PCR. Forty-two (36.5%) samples were positive for isolated L452R mutation, and were presumptive B.1.427/B.1.429 variant. In unadjusted analysis, PVSCs infected with L452R-containing viruses were more likely than those with no identified mutations to have tested positive after the second vaccine dose, or to be partially- or fully-vaccinated at time of positive test (Table 1); this variant was not associated with age, gender, job role, brand of vaccine received, immunocompromised status, or symptomatic infection. In multivariate analysis, when controlling for community prevalence of L452R mutation the week prior to positive test (when exposure likely occurred), vaccination status at time of positive test was not significantly associated with presumptive B.1.427/B.1.429 (Table 3).

**Table 3.** Risk ratios for infection with presumptive B.1.427/B.1.429 among PVSC by vaccination status and adjusted for community prevalence of L452R mutation at time of infection.

| Vaccination status at time of positive test | n | L452R Community Prevalence Median (quartiles) | Presumptive B.1.427/B.1.429 n (%) | Unadjusted RR (95% CI) | Adjusted RR (95% CI) |
| --- | --- | --- | --- | --- | --- |
| **Early post-vaccination** | 74 | 36.2% (32.8%, 41.4%) | 22 (29.7%) | Ref. | Ref. |
| **Partially Vaccinated** | 21 | 41.4% (41.4%, 44.1%) | 10 (47.6%) | 1.61 (0.90-2.84) | 1.40 (0.81-2.43) |
| **Fully Vaccinated** | 17 | 51.8% (50.7%, 57.7%) | 10 (58.8%) | 1.98 (1.16-3.37) | 1.13 (0.59-2.16) |

Modified Poisson regression model (log-Poisson generalized linear model with robust standard errors) comparing individuals with and without presumptive B.1.427/B.1.429 by vaccination status at time of positive SARS-CoV-2 NAAT and adjusted for percent of total positive SARS-CoV-2 tests with isolated L452R mutation presumptive of B.1.427/B.1.429 within Stanford Health Care during week of infection. Early post-vaccination = Positive SARS-CoV-2 NAAT $\leq 14$ days from vaccine dose 1; Partially vaccinated = Positive SARS-CoV-2 NAAT $> 14$ days from vaccine dose 1 and $\leq 14$ days from vaccine dose 2; Fully vaccinated = Positive SARS-CoV-2 NAAT $> 14$ days from vaccine dose 2.

## Discussion

Here we present descriptive data on HCP who tested positive for SARS-CoV-2 after receiving at least one dose of either Pfizer or Moderna mRNA-based SARS-CoV-2 vaccine. As others have reported(5, 6), we found that positive tests post-vaccination are uncommon, with only 189 such individuals identified out of a total of >22,000 vaccinated people despite widespread community circulation.(25) Further, most of these positive tests occurred in the first 2 weeks after vaccination, before immunity is expected to develop. It is unclear whether these individuals were exposed before or after their vaccine dose, though the reported onset of symptoms prior to first vaccine dose among some individuals, and the typical incubation period of 5-9 days(26) suggests that several of these early post-vaccination infections were acquired prior to the first vaccine dose. Twenty-six PVSCs occurred >14 days after the second dose, when full immunity is expected. Although most infections were symptomatic, only 2 individuals required hospitalization; both developed symptoms and tested positive within 14 days of the first vaccine dose. These findings are consistent with other real-world reports of excellent vaccine effectiveness >14 days after the first and second doses(5, 6, 27) particularly in preventing severe disease, and highlights the need to observe strict precautions until full immunity is achieved >14 days after the second vaccine dose.

Isolated L452R mutation presumptive of infection with B.1.427/B.1.429 variant was associated with partially- and fully-vaccinated status on univariate analysis. This finding is likely attributable to the timing of vaccinations (as most SHC HCPs were first vaccinated in December 2020), coincident with the rising overall prevalence of B.1.427/B.1.429 from December 2020 to March 2021, rather than to reduced vaccine effectiveness against B.1.427/B.1.429. However, on multivariate analysis controlling for overall community prevalence of the variant, infection with presumptive B.1.427/B.1.429 was not significantly more common in partially-and fully-vaccinated PVSCs than in early PVSC. Although we cannot conclude that vaccinated individuals are more at risk of infection with B.1.427/B.1.429 compared with wild-type SARS-CoV-2 based on this small cohort, we believe further vigilance is warranted. Reduced in vitro neutralizing activity of naturally- and vaccine-acquired antibodies against the B.1.427/B.1.429 compared with wild-type S protein has been reported.(10, 16) As more people become fully vaccinated, careful surveillance will be needed to determine if variants such as B.1.427/B.1.429 can escape vaccine-derived protection in vivo, and continued caution is needed especially as states begin to relax restrictions intended to curb transmission.

Only three PVSCs with N501Y mutation alone were observed, and no E484K mutations were found. The PVSCs with N501Y mutation occurred towards the end of the project in late March 2021. N501Y in the absence of E484K could indicate B.1.1.7 (the UK variant), which is expected to be the dominant variant in the United States in Spring 2021.(28) In this cohort of PVSCs in HCP, L452R was much more prevalent.

Higher C_t_ value, which corresponds with lower viral load, in individuals testing positive >14 days after first vaccine dose suggests some protection from viral replication and reduction in transmissibility in people who contract SARS-CoV-2 post vaccination. This finding is consistent with reports of real world experience in mass vaccination campaign in Israel.(29) Lower C_t_ value, corresponding with higher viral load, was seen in vaccinated individuals who reported symptoms consistent with COVID-19 compared with the few who remained asymptomatic.

The peak we observed in post-vaccine positive SARS-CoV-2 cases in late December 2020 to early January 2021 corresponds with a rise in county-wide cases and HCP cases at that time. We did not find any evidence to support transmission clusters or superspreading events contributing to post-vaccination SARS-CoV-2 infections within the health care system. Although several PVSCs had been vaccinated on the same two days in December, nearly half of these infections occurred more than two weeks after vaccination, suggesting that exposure occurred sometime after the date of vaccination. Further, 41.8% reported household contacts with known SARS-CoV-2 infection at the time of their positive test, suggesting community transmission was driving these cases rather than workplace exposure; the multiple cases in late December and early January may have been related to holiday celebrations. This highlights the importance of maintaining social distancing and diligent mask wearing at work in the setting of increased prevalence of variants. Describing the phenomenon of PVSCs in this quality improvement project has allowed us to more effectively promote these measures within our system at a time when many are tempted to loosen precautions due to “pandemic fatigue” and a sense of security after being vaccinated.

We do note several limitations to this report. We used time of test in relation to vaccination (before or after 14 days post-vaccination) as the comparator, assuming that early positive tests were prior to when vaccine is expected to afford protection; however, other than county data, we did not have a comparison group of unvaccinated individuals that had similarly high levels of surveillance and detailed demographic and clinical information available. Some employees may also have been tested outside the SHC system; although employees are required to report such positive tests, we cannot be certain that all PVSCs were captured. Though we report total number of tests and number of positive tests performed in the SHC system among employees during this time period, this may include repeated testing of some individuals and therefore is not a perfectly accurate denominator. Nevertheless, we believe our findings are an important contribution to the understanding of PVSCs in the context of rising prevalence of new SARS-CoV-2 variants.

## Conclusion

The great majority of 189 PVSCs, as expected, occurred prior to full, vaccine-derived immunity. Presumptive B.1.427/B.1.429 did not represent a significantly higher proportion of late PVSCs than would be expected based on background variant incidence but numbers of PVSCs were small. Continued infection control measures in the workplace and in the community including social distancing and masking, particularly in the early days post-vaccination, as well as continued variant surveillance in PVSCs, is imperative in order to anticipate and control future surges of infection.

## Data Availability

Data is available upon request.

## Acknowledgments

The authors would like to thank Stanford Health Care’s occupational health team, as well as all SHC healthcare personnel.

